# Sustainable social distancing through facemask use and testing during the Covid-19 pandemic

**DOI:** 10.1101/2020.04.01.20049981

**Authors:** Diego Chowell, Kimberlyn Roosa, Ranu Dhillon, Gerardo Chowell, Devabhaktuni Srikrishna

## Abstract

We investigate how individual protective behaviors, different levels of testing, and isolation influence the transmission and control of the COVID-19 pandemic. Based on an SEIR-type model incorporating asymptomatic but infectious individuals (40%), we show that the pandemic may be readily controllable through a combination of testing, treatment if necessary, and self-isolation after testing positive (TTI) of symptomatic individuals together with social protection (e.g., facemask use, handwashing). When the basic reproduction number, R0, is 2.4, 65% effective social protection alone (35% of the unprotected transmission) brings the R below 1. Alternatively, 20% effective social protection brings the reproduction number below 1.0 so long as 75% of the symptomatic population is covered by TTI within 12 hours of symptom onset. Even with 20% effective social protection, TTI of 1 in 4 symptomatic individuals can substantially 'flatten the curve' cutting the peak daily incidence in half.

---

Recent reports have documented substantial proportions of asymptomatic or mildly symptomatic Covid-19 infections that may also be infectious [1–3] as well as the potential role of respiratory droplets and contaminated surfaces in driving SARS-CoV-2 transmission [4]. While facemask use is promoted in healthcare settings as part of infection control protocols [5–8], its use during the COVID-19 pandemic varies across countries [9,10]. As the COVID-19 pandemic rages on, widespread facemask use has been recently recommended by Dr. George Gao, the director-general of the Chinese Center for Disease Control and Prevention (CDC) [11].

Any plan for stopping the ongoing 2019-nCov pandemic must be based on a quantitative understanding of the proportion of the population that needs to be protected by effective control measures such that each infected person infects no more than one other person on average (the effective reproductive number, R < 1), at which point transmission contracts and eventually burns out. Based on a modeling analysis, we show that the pandemic may be readily controllable through a combination of testing, treatment if necessary, and self-isolation after testing positive (TTI) of symptomatic individuals together with social protection (e.g., facemask use, handwashing).

We used an SEIR-type model incorporating asymptomatic but infectious individuals (40%) [1–3] to investigate how individual protective behaviors, different levels of testing, and isolation influence the transmission and control of the COVID-19 pandemic (Appendix). When the basic reproduction number, R_0_, is 2.4 [12], 65% effective social protection alone (35% of the unprotected transmission) brings the R below 1 (Appendix). Alternatively, 20% effective social protection brings the reproduction number below 1.0 so long as 75% of the symptomatic population is covered by TTI within 12 hours of symptom onset (Fig. 1A and Appendix). Even with 20% effective social protection, TTI of 1 in 4 symptomatic individuals can substantially “flatten the curve”, cutting the peak daily incidence in half (Fig. 1B and Appendix).

Therefore, our results suggest that the COVID-19 pandemic can be controlled through an imperfect though sufficient combination of testing, contact tracing, and protective measures like masks and distancing when in public.

**Figure 1.**
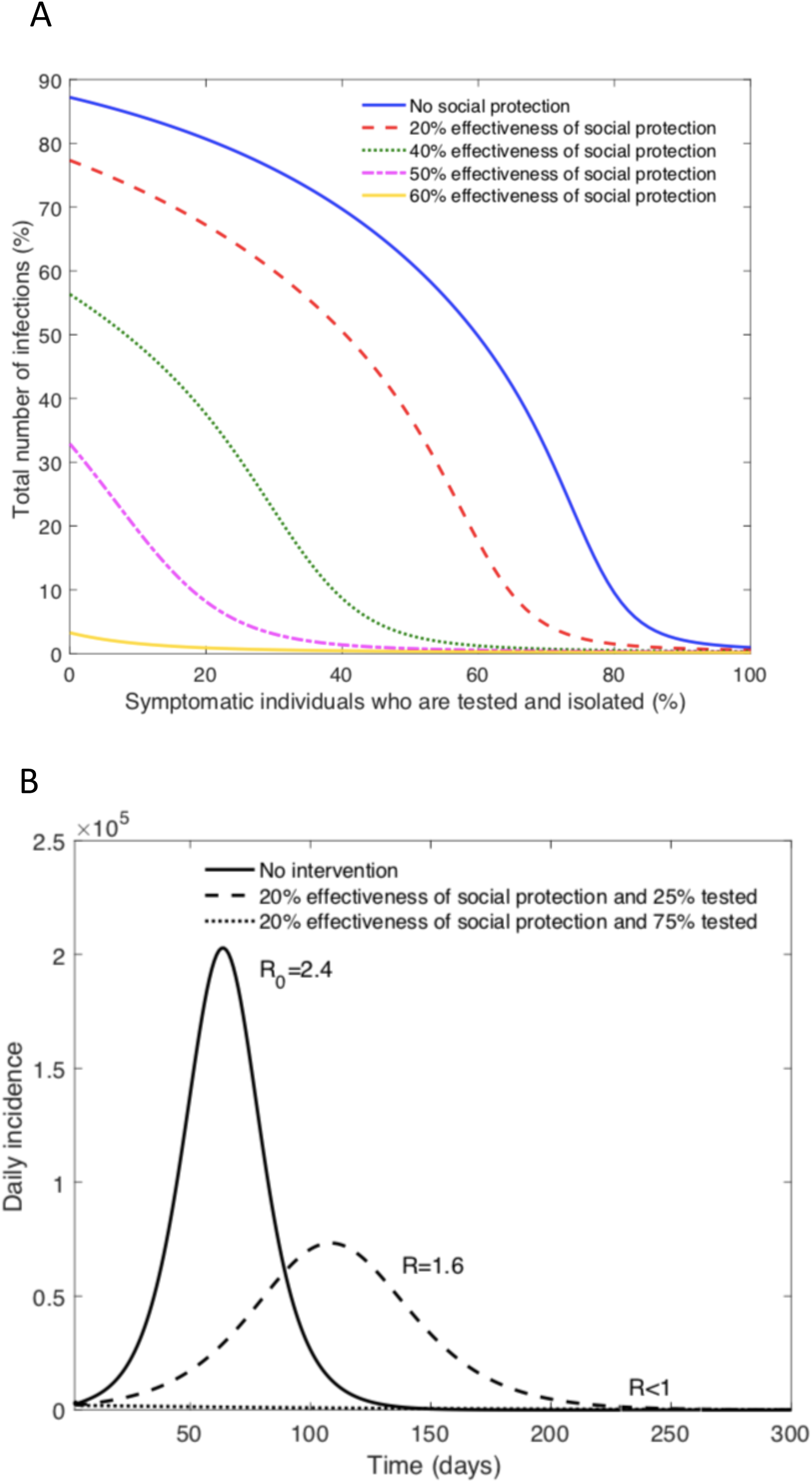
(A) Total number of infections in a population of 10 million individuals for varying levels of social protection and testing (TTI) at 12 hours testing delay and assuming 40% asymptomatic transmission. (B) Daily incidence in a population of 10 million individuals for 20% effectiveness of social protection and varying levels of testing (TTI) at 12 hours testing delay and assuming 40% asymptomatic transmission. Initial conditions were set as follows, susceptible individuals: 9,943,400; symptomatic individuals who underwent testing: 40,000; asymptomatic individuals: 16,000; and deceased individuals: 600.

## Data Availability

All data is included in the appendix

## Appendix Sustainable social distancing through facemask use and testing during the Covid-19 pandemic

### Basic reproduction number, R_0_

The basic reproduction number, R_0_, is defined as the average number of secondary cases generated by primary infectious individuals during the early transmission phase in a completely susceptible population and in the absence of control interventions. This is a key metric to gauge the intensity and type of interventions that need to be implemented in order to bring the epidemic under control. For the ongoing COVID-19 pandemic, R_0_ has been estimated at 2.4 [1].

### Reproduction number with testing, isolation, and social protection

The reproductive number, R, quantifies the potential for infectious disease transmission in the context of a partially susceptible population. When R > 1, infection may spread in the population, and the rate of spread is higher with increasingly high values of R. If R < 1, infection cannot be sustained and is unable to generate an epidemic. As there are seven classes that can contribute to new infections, the reproduction number is the sum of the contributions of the infectious classes:

The contributions of the individual compartments are as follows

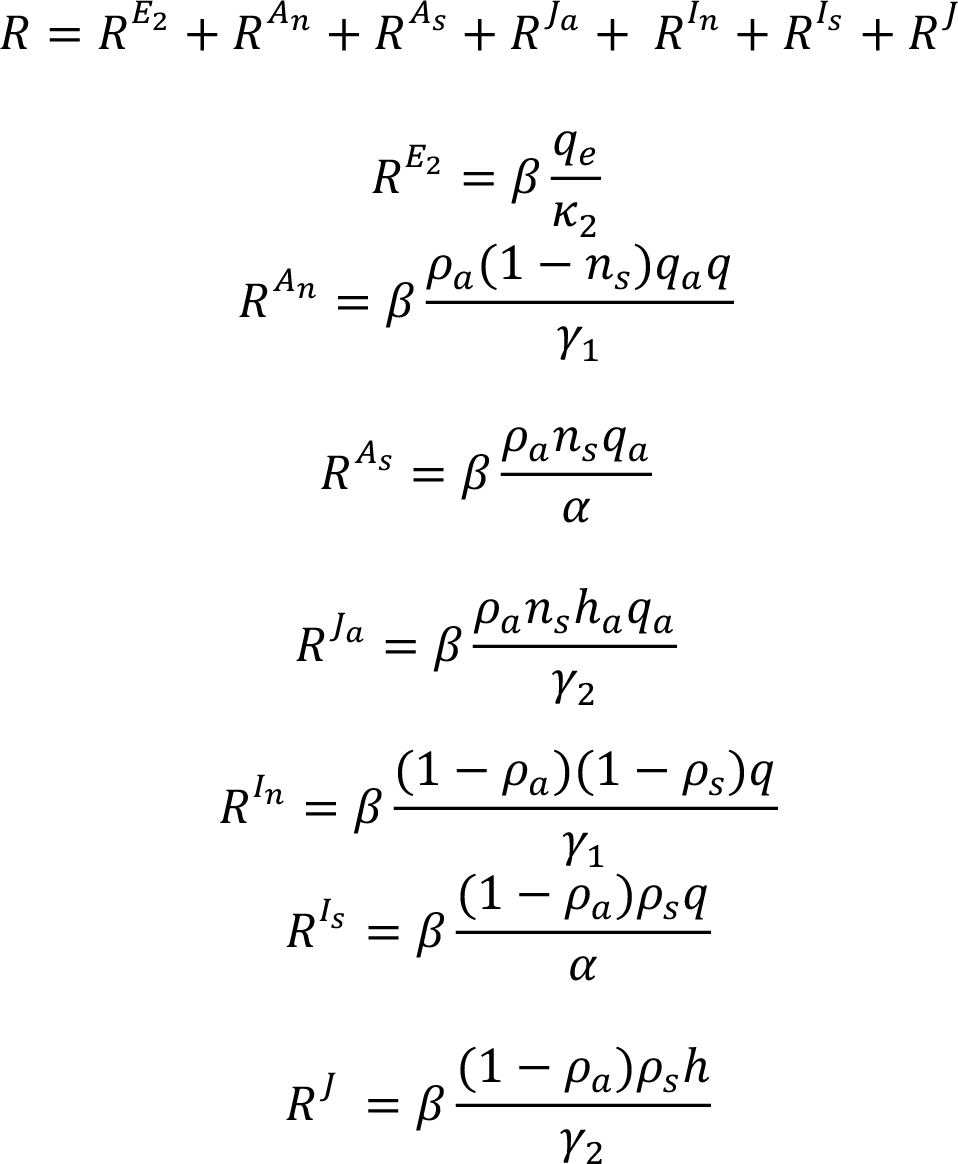

### Modeling the effect of testing, isolation and social protection of infected individuals

Compartmental transmission models are commonly used in infectious disease epidemiology as a population-level modeling approach that subdivides the population into various classes based on the epidemiological status. Compartmental models are specified by a set parameters and ordinary differential equations that track the progression of the number of individuals within each class. Here, we developed an extension of the commonly used SEIR transmission model for modeling the transmission of COVID-19.

Individuals within the model are classified as susceptible (S), latent (E_1_), partially infectious but not yet symptomatic (E_2_), asymptomatic who are not tested (A_n_), asymptomatic who are tested (A_s_), infectious symptomatic who are not tested (I_n_), infectious symptomatic who are tested (I_s_), isolated infectious symptomatic (J), isolated asymptomatic (J_a_), recovered (R), and deceased (D). Constant population size is assumed, so *N* is equal to the sum of individuals in all of the compartments. Further, seven classes can contribute to new infections: E_2_, A_n_, A_s_, I_n_, I_s_, J, J_a_. Susceptible individuals move to the latent E_1_ class at a rate *β[q_e_ E*_2_(*t*) + *q_a_ qA_n_*(*t*) + *q_a_A_s_*(*t*) *+ qI_n_*(*t*) + *qI_s_*(*t*) + *h_a_J_a_*(*t*) + *hJ*(*t*)*]/N,* where β denotes the transmission rate. The transmission rate, β, was calibrated based on the baseline value of the basic reproductive number, R_0_ = 2.4 [1]. Parameter *q_e_* represents the relative transmissibility of exposed individuals in E_2_; *q_a_* denotes the relative transmissibility of asymptomatic individuals; *q* denotes the level of effectiveness of social protection such as wearing facemasks. *h* represents the relative transmissibility of symptomatic individuals in isolation; *h_a_* denotes the relative transmissibility of asymptomatic individuals in isolation.

Individuals in E_1_ progress to E_2_ at rate κ_1_. Individuals from E_2_ are partially infectious, with relative transmissibility q_e_, and progress at a rate κ_2_, where a proportion pa become asymptomatic and partially infectious (relative transmissibility q_a_), and 1 − ρ_a_ become infectious. Among the proportion *ρ*_a_ who become asymptomatic, ns are tested, while 1 - n_s_ are undetected. Further, among the proportion 1 − ρ_a_ that become fully infectious, ρ_s_ are tested, while 1 − ρ_s_ will be undetected. Asymptomatic individuals who are not tested and symptomatic individuals practice social protective behaviors such as wearing masks in public and increased handwashing, and thus have relative transmissibility q, which quantifies the effectiveness of those protective behaviors. Individuals within A_n_ and I_n_ classes (individuals who are not tested) recover at rate γ_1_. Those who are tested (I_s_ and A_s_) progress to the isolation class at diagnosis rate α. Symptomatic individuals who are in isolation have a relative transmissibility *h*. Also, asymptomatic individuals in isolation have a relative transmissibility *h_a_.* However, we assume perfection isolation for simplicity (i.e. *h=0* and *h_a_=0).* We also assume that asymptomatic individuals are not detected. Individuals who are isolated progress to the recovered class at a rate *γ*_2_ or to the deceased class at a rate *δ*.

Therefore, the system is defined by the following system of non-linear differential equations:

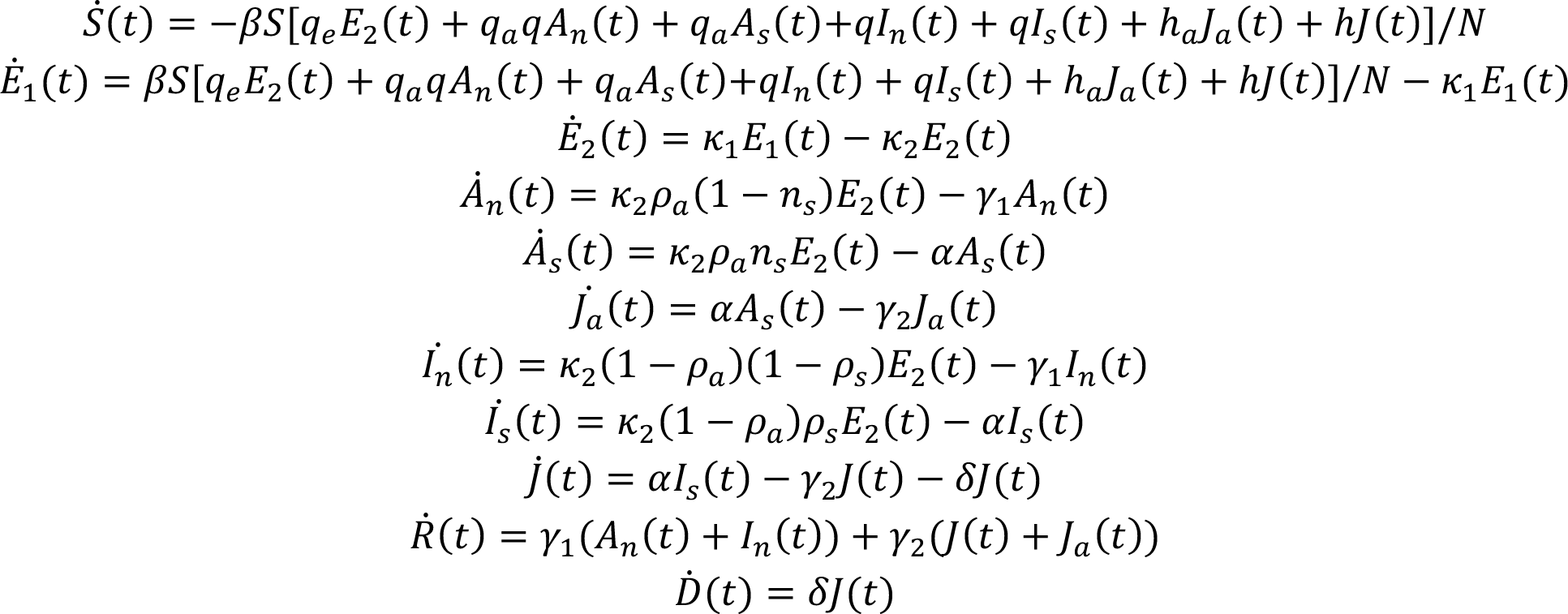

### Schematic of the model diagram

The population is classified into 11 epidemiological states: susceptible (S), latent (E_1_), partially infectious but not yet symptomatic (E_2_), asymptomatic who are not tested (A_n_), asymptomatic who are tested (A_s_), infectious symptomatic who are not tested (I_n_), infectious symptomatic who are tested (I_s_), isolated infectious symptomatic (J), isolated asymptomatic (J_a_), recovered (R), and deceased (D). Model parameters are described in Table 1.

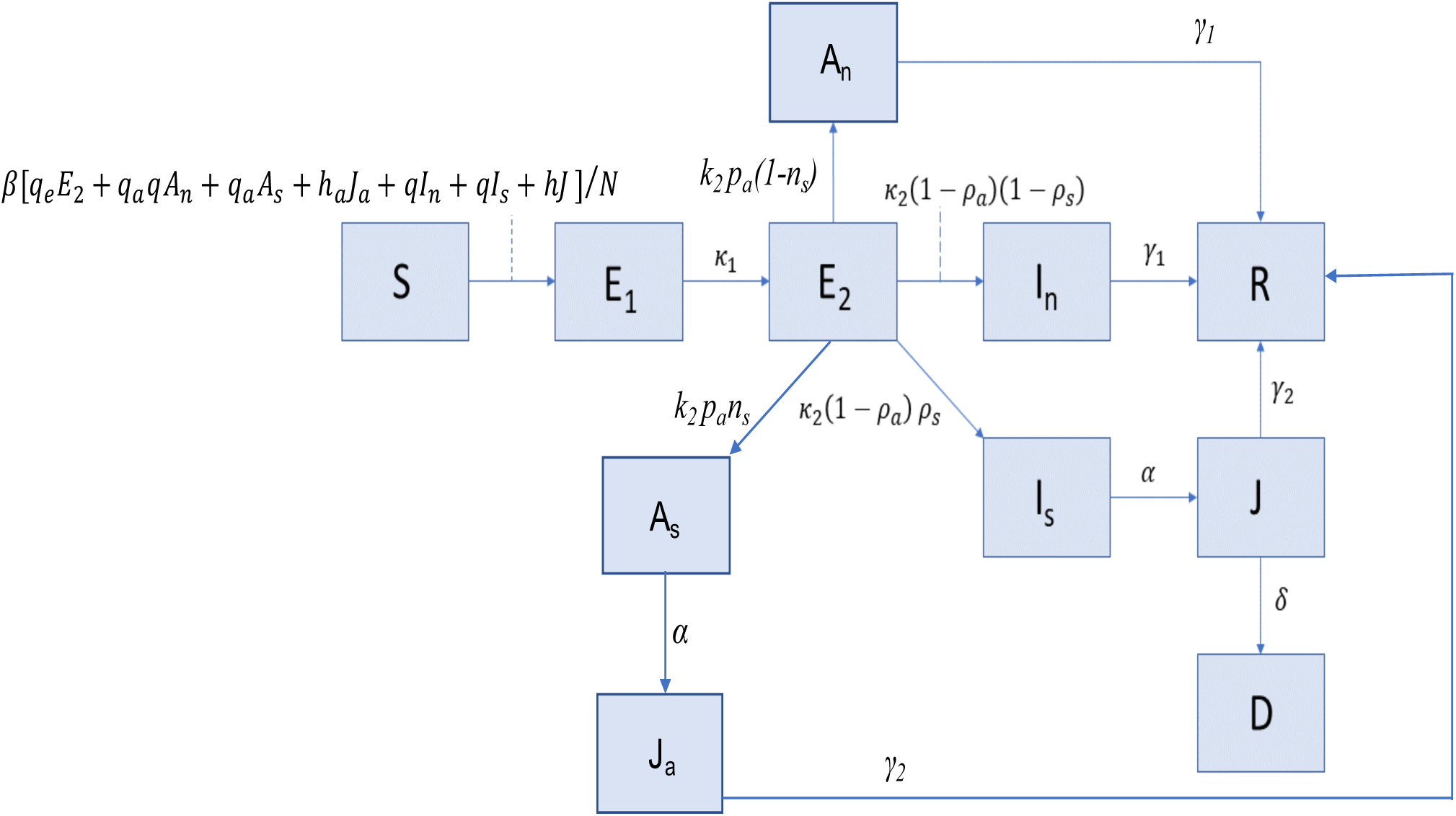

**Table 1.** Parameter descriptions and values for model

| Parameter | Description | Value | References |
| --- | --- | --- | --- |
| $\beta$ | Transmission rate | Calibrated for $R_0 = 2.4$ | ** |
| $h$ | Relative transmissibility of symptomatic individuals in isolation | 0-1 | Assumed here |
| $h_a$ | Relative transmissibility of asymptomatic individuals in isolation | 0-1 | Assumed here |
| $q_e$ | Relative transmissibility of exposed individuals in $E_2$ | 0.1 | [2] |
| $q_a$ | Relative transmissibility of asymptomatic individuals | 0.4 | [2] |
| $q$ | Level of effectiveness of personal protective behavior | 0-1 | |
| $1/\kappa_1$ | Mean latency period in $E_1$ | 2.5 days | [3-7] |
| $1/\kappa_2$ | Mean latency period in $E_2$ | 2.5 days | [5-8] |
| $\rho_a$ | Proportion of asymptomatic infections | 0.2, 0.4, and 0.6 | [9, 10] |
| $n_s$ | Proportion of asymptomatic individuals who are tested | 0-1 | Assumed here |
| $\rho_s$ | Proportion of symptomatic infectious individuals that undergo testing | 0-1 | |
| $1/\alpha$ | Time from detection to confirmation and isolation | 1-2 days, and 12 hours | |
| $1/\gamma_1$ | Recovery rate for individuals in $A_n$ or $I_n$ | 7 days | [3, 4] |
| $1/\gamma_2$ | Average time from confirmation to recovery | 5 days | [11] |
| $\delta$ | Disease-induced death rate among confirmed symptomatic individuals | $\delta = 0.04$ | [12] |

\*\*Indicates the value was estimated

**Figure 1.**
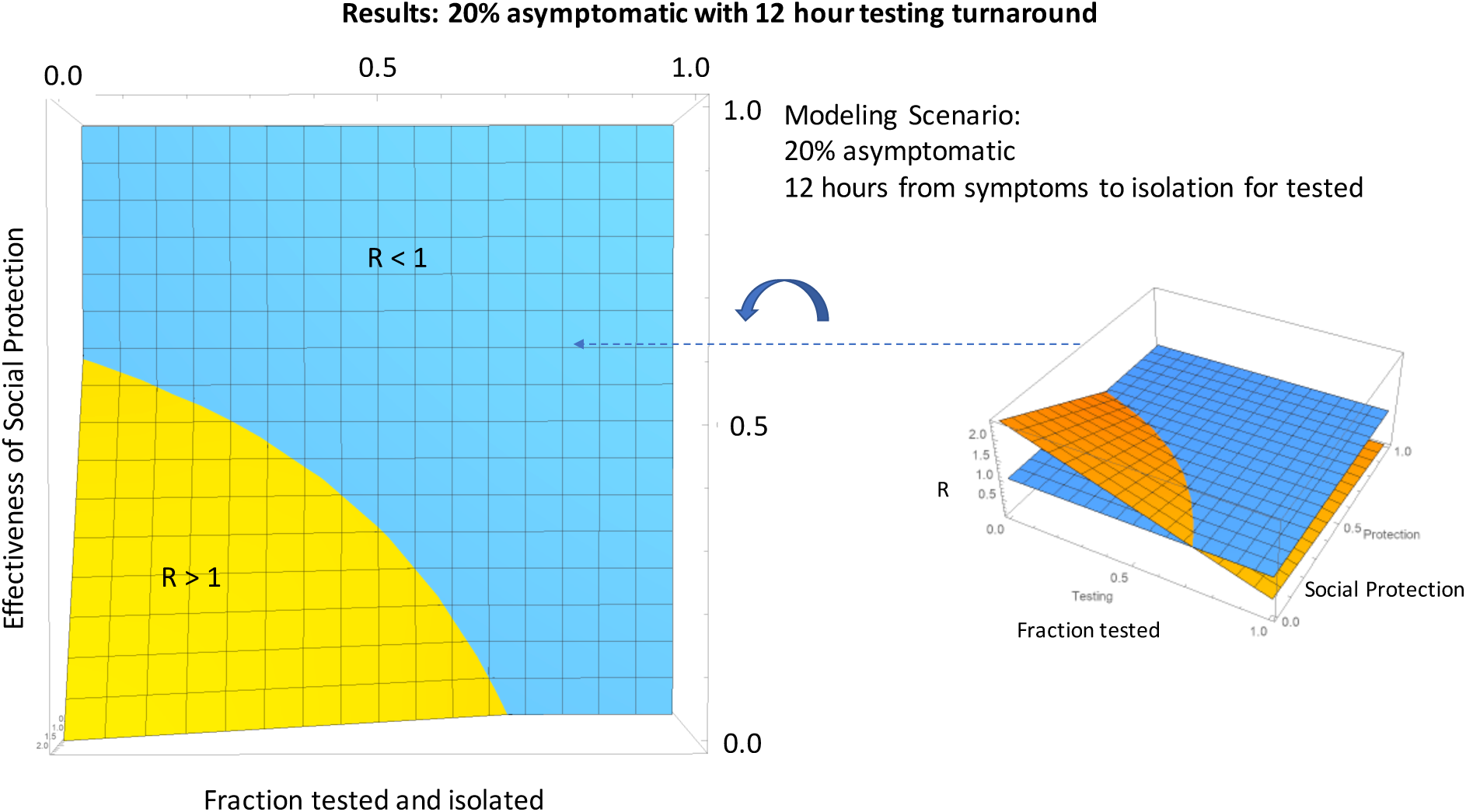
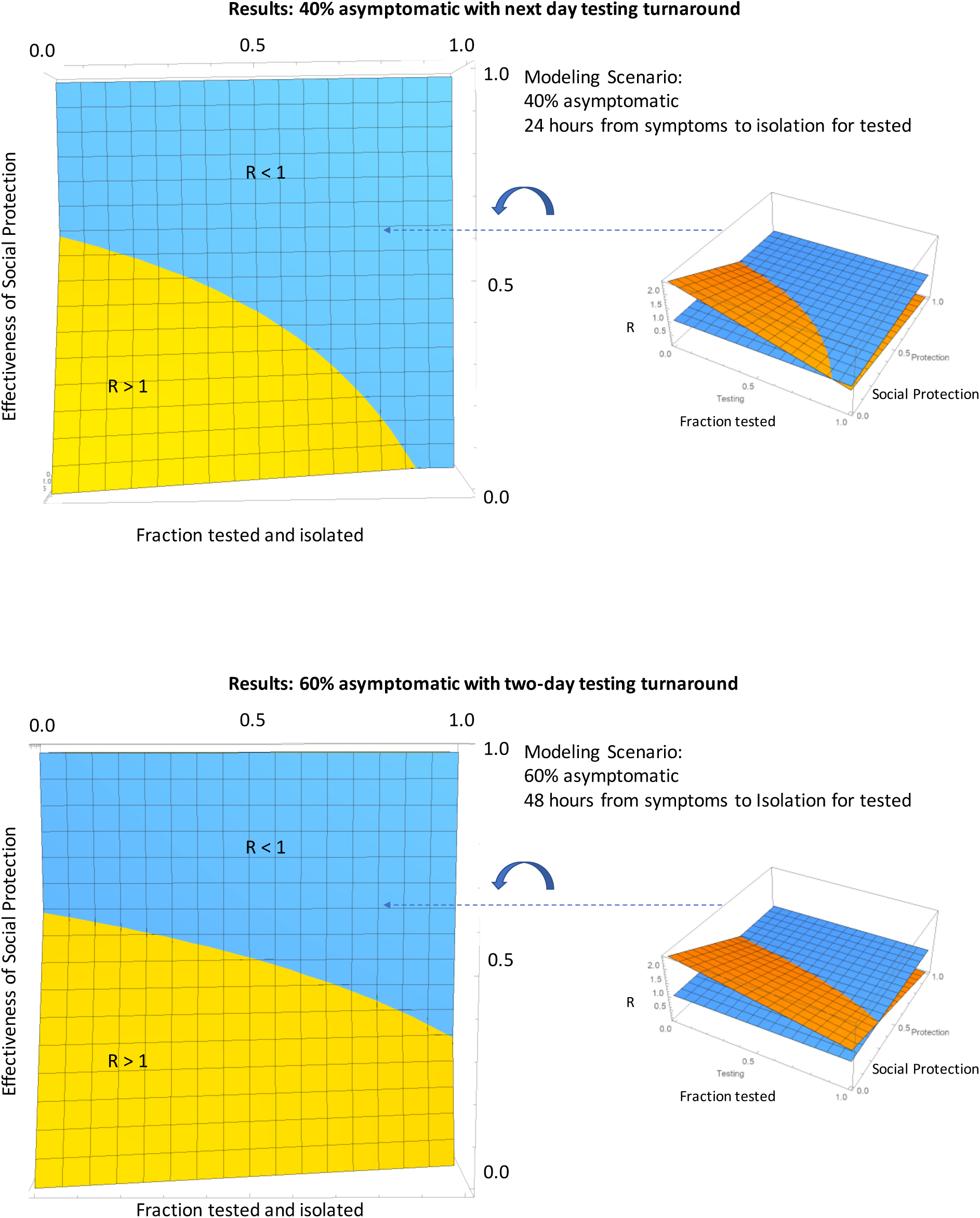
Thresholds for social protection and testing / isolation needed to bring R < 1.

## Notes

### Competing Interest Statement

The authors have declared no competing interest.

### Funding Statement

none

